# Expression of FIBCD1 by intestinal epithelial cells alleviates inflammation-driven tumorigenesis in a mouse model of colorectal cancer

**DOI:** 10.1101/2023.08.15.23293749

**Authors:** Vahid Khaze Shahgoli, Magdalena Dubik, Bartosz Pilecki, Sofie Skallerup, Sandra Gaedt Schmidt, Sönke Detlefsen, Grith L. Sorensen, Uffe Holmskov, Behzad Baradaran, Jesper B. Moeller

## Abstract

Colorectal cancer (CRC) ranks as the third most prevalent cancer worldwide, highlighting the urgent need to address its development. Inflammation plays a crucial role in augmenting the risk of CRC development and actively contributes to all stages of tumorigenesis. Consequently, targeting early inflammatory responses in the intestinal tract to restore homeostasis holds significant potential for innovative therapeutic strategies against CRC. In this study, we employ transgenic mice that mimic human expression of Fibrinogen C domain containing 1 (FIBCD1), a chitin-binding transmembrane protein primarily present on intestinal epithelial cells (IECs). Previous research has highlighted FIBCD1’s ability to effectively suppress inflammatory responses and foster tissue homeostasis at mucosal barriers. Using the azoxymethane/dextran sodium sulfate mouse model, we demonstrate that FIBCD1 substantially impacts CRC development by significantly reducing intestinal inflammation and suppressing colorectal tumorigenesis. Moreover, we identify a soluble variant of FIBCD1, which is significantly increased in fecal matter during acute inflammation. Together, these findings suggest that FIBCD1 has the potential to be a novel molecular target in the context of colitis-associated colorectal cancer and emerges as an intriguing candidate for future research.

## 1. Introduction

Colorectal cancer (CRC) is the third leading cause of cancer-related deaths with an incidence of more than 1.8 million annual cases [1]. It is well established that inflammation plays a prominent role in tumorigenesis and the progression of several different types of cancers, including CRC [2-4]. Prolonged intestinal inflammation observed in patients with inflammatory bowel disease (IBD) greatly increases the risk of CRC development, yet the molecular pathogenesis of colitis-associated colorectal cancer (CAC) remains poorly defined [5]. It has been suggested that repeated cycles of cell damage as well as sustained production of reactive nitrogen and oxygen species at the site of inflammation can damage cellular DNA, and thereby favor the formation of malignant cells [6]. Furthermore, the inflammatory microenvironment exhibits tumor-promoting effects through the production of inflammatory mediators that can drive cancer cell proliferation, aid metastasis, and influence the efficiency of chemotherapeutic drugs [2]. In contrast to CRC, which generally arises from adenomatous polyps and mainly occurs in older patients, CAC is more frequently diagnosed in younger patients and arises from flat dysplastic mucosa as a consequence of chronic inflammation [7, 8].

Animal models of CAC, such as the azoxymethane (AOM)/dextran sodium sulfate (DSS) mouse model, have emerged as important tools that mimic aspects of the human disease and improve our understanding of the underlying pathological mechanisms [9]. The AOM/DSS model is a two-step tumor model of CAC in which administration of AOM, a pro-carcinogen that causes DNA damage in colonic epithelial cells, is followed by repeated exposure to DSS that inflicts mucosal damage leading to chronic colitis as well as the development of distally located invasive adenocarcinomas [9-11]. The AOM/DSS model effectively recapitulates the CAC development from inflammation to malignancy, allowing the investigation of pathways and novel target molecules that can lead to improved treatment strategies.

Fibrinogen C domain containing 1 (FIBCD1) is a membrane protein expressed in human epithelial cells at mucosal barrier surfaces, such as the gastrointestinal and respiratory tract [12]. Acting as a pattern recognition receptor, FIBCD1 specifically recognizes acetylated compounds such as chitin, the structural component of e.g., invertebrates, insects, and fungi [13]. Previous studies have shown that FIBCD1 expression in epithelial cells can ameliorate fungal-driven intestinal inflammation and promote tissue homeostasis [14, 15]. Moreover, studies have demonstrated that FIBCD1 expression in lung epithelium can influence inflammatory responses to fungal pathogens [16, 17]. Based on FIBCD1’s ability to dampen inflammatory responses in mucosal tissues, we hypothesized that enhanced FIBCD1 expression would lead to decreased intestinal inflammation and reduce the development of colitis-associated colon cancer. By employing transgenic mice that specifically express FIBCD1 in the intestinal epithelium and subjecting them to AOM/DSS treatment we demonstrate that increased FIBCD1 expression ameliorates the development of CAC.

## 2. Materials and Methods

### 2.1. Mice

*Fibcd1*-transgenic mice (TG) overexpressing *Fibcd1* in intestinal epithelial cells (IEC) and wild-type littermates (WT) C57BL/6 mice were generated and maintained under standard conditions (12-h light/dark cycle, 21–24 °C, and 55% relative humidity) at the Biomedical Laboratory, University of Southern Denmark [14]. All experiments were performed using co-housed, sex- and age-matched littermates and conducted in accordance with the national ethical committee (Animal Experiments Inspectorate under Danish Ministry of Food, Agriculture and Fisheries, The Danish Veterinary and Food Administration, approval identification number: 2019-15-0201-00349).

### 2.2. AOM / DSS model

Colitis-associated colorectal cancer (CAC) was induced in mice by combinatory treatment with azoxymethane (AOM) and dextran sulfate sodium (DSS) as previously described [2]. In brief, TG and WT littermates were randomized to receive a single intraperitoneal (i.p.) injection with AOM (10 mg/kg body weight, Sigma Aldrich). 3 days later, chronic colitis was induced by three cycles of 1.5% DSS (molecular weight 36,000– 50,000 Da, Colitis Grade, MP Biomedicals) in drinking water for 5-7 days followed by two weeks of recovery with normal drinking water (Figure 2A). Mice were monitored daily for weight and general health. Due to ethical considerations, mice were sacrificed if weight loss exceeded 20% of the initial body weight or if they showed significant signs of pain or distress.

### 2.3. Tissue collection and processing

Mice were euthanized by cervical dislocation 15 days after the final DSS treatment. The entire large intestines were harvested and measured (Figure 2C). Afterward, the colons were separated, flushed with phosphate-buffered saline (PBS), cut open longitudinally, and spread on a filter paper with the intestinal lining facing up. The tissue specimens were stained with methylene blue (2% in PBS) for 1 minute, washed in PBS, and photographed for tumor counting and measuring. A piece of the tumor and the adjacent tissue were taken from each colon for RNA extraction. For histopathological analysis, the remaining tissues were destained with fresh PBS, swiss-rolled, and fixed in 10% buffered formalin (Sigma) for 48 h. The tissues were then embedded in paraffin, sectioned into 4 μm thick slides, and stained with hematoxylin and eosin (H&E).

### 2.4. RNA isolation and cDNA synthesis

RNA was extracted from colon tissue samples using TRI Reagent® (Sigma-Aldrich) according to the manufacturer’s instructions. For tissues isolated from DSS-treated mice, a lithium chloride (8 M LiCl) precipitation step was performed to remove DSS contamination from the RNA [18]. RNA concentration was determined using a NanoDrop™ One spectrophotometer (Thermo Fisher Scientific) and the total RNA was converted into cDNA using the High-Capacity cDNA Reverse Transcription kit (Thermo Fisher Scientific) according to the manufacturer’s instructions.

### 2.5. Fecal DNA purification

Fecal samples were collected from WT and TG littermates at eight different time points: Before AOM treatment (day -3), at the initiation of DSS treatment (day 0), and on days 10, 22, 32, 44, 54, and 65 after AOM treatment. One or two fecal pellets from each mouse were collected, snap-frozen in liquid nitrogen, and stored at −80°C until further processing. Before DNA extraction, the samples were treated with lyticase (Sigma-Aldrich) for 1 h at 30°C, followed by bead-beating, and isolation using the QIAamp® Fast DNA Stool kit (Qiagen) as described previously [14].

### 2.6. RT-qPCR and qPCR

Quantification of gene expression from colonic and tumor tissues was performed using the TaqMan Gene Expression Assays (Applied Biosystems) and the SYBR green primers listed in Supplementary Table 1. The data were generated on a StepOnePlus™ Real-Time PCR (Thermo Fisher Scientific) and the results were normalized to *Hprt1* using the 2–ΔCt method. For fecal microbiota analysis, the SYBR green primers listed in Supplementary Table 2 were used. Relative abundance was standardized to input DNA using 16S as a reference. To counteract DSS inhibition of PCRs, the samples were supplemented with 0.01 g/l spermine (Sigma-Aldrich) right before analysis [19].

### 2.7. Fecal Inflammatory Biomarkers

Fecal samples were suspended in PBS containing 2 mM EDTA to a final concentration of 200 mg/ml. Subsequently, the samples were homogenized, vortexed vigorously, and the supernatants were collected by centrifugation at 10.000 g for 10 min. Fecal lipocalin (LCN2) and calprotectin (CAL) levels were determined using enzyme-linked immunosorbent assays (ELISAs): Mouse Lipocalin-2/NGAL DuoSet® ELISA kit (Cat no. DY1857-05) and Calprotectin (Cat no. DY8598-05) (R&D systems, Minneapolis, MN, USA) according to the manufacturer’s recommendations. For the detection of soluble FIBCD1 in fecal specimens a sandwich ELISA was developed using monoclonal antibodies raised against the fibrinogen-related domain of FIBCD1 previously produced [14]. In brief, MaxiSorp 96-well plates (NUNC) were coated with 1µg/ml of monoclonal anti-FIBCD1 antibody (clone 11-14-40) in standard ELISA coating buffer (pH 9.5), followed by blocking in 5% BSA in PBS. PBS supplemented with 0.05% (vol/vol) Tween20 was used for all washing steps. Samples were loaded in diluent buffer (10 to 10,000-fold, diluted in PBS/1% BSA) and compared to a recombinant FIBCD1 standard. Soluble FIBCD1 and standards were detected using biotinylated 1 µg/ml of monoclonal anti-FIBCD1 antibody (clone 11-14-25) and quantified using streptavidin/HRP combined with TMB substrate according to conventional ELISA methods.

### 2.8. Histopathological score

Intestinal inflammation was assessed in a blinded fashion using a previously established protocol [20]. Briefly, colonic sections were stained with H&E and scanned using the slide scanner NanoZoomer XR (HAMAMATSU, Japan). The sections were evaluated for disease severity based on scoring of inflammatory cell infiltration (graded 0-3) and overall colonic architecture (graded 0-3). The sum of the two values gave a total colitis severity score between 0 and 6. The analysis was performed using the NDP.view2 software (HAMAMATSU, Japan). Tumor burden and volume were analyzed using ImageJ v1.49 software.

### 2.9. Human surgical CRC specimens and FIBCD1 immunoscore

35 formalin-fixed paraffin-embedded tissue specimens prepared from surgically removed CRC patient material as well as information about disease stage were obtained from the Immunology Research Center, Tabriz University of Medical Sciences. Staining for FIBCD1 was performed on 4 µm tissue sections as previously described [12]. The semiquantitative immune score for FIBCD1 expression was performed using a scoring system consisting of a distribution score and an intensity score, as described previously [21]. The FIBCD1 positive epithelial cell distribution was scored from 0 to 4 as followed: 0, less than 10% of the epithelial cells were positive for FIBCD1; 1, 10%–30% of the epithelial cells were positive for FIBCD1; 2, 31%–60%, 3, 61-90%, and 4, >90% of the epithelial cells were positive for FIBCD1. The intensity score was graded as 0, no expression; 1, weak expression; 2, moderate expression; and 3, strong expression. The sum of the distribution score and the intensity score composed the FIBCD1 immunoscore (0-7). All immunostainings were evaluated by a trained gastrointestinal pathologist, blinded to clinical data.

### 2.10. Statistical analysis

Statistical analysis was performed using GraphPad Prism v9.4.0 (GraphPad Software Inc, San Diego, California USA). Student’s t-test with Welch’s correction for unequal SD was used to compare single parameters between two datasets. Single parameters with more than two datasets were analyzed using one-way ANOVA followed by the Tukey multiple comparisons test. Weight curves were analyzed by a mixed-effects model (REML) of two-way repeated measures ANOVA followed by Holm-Šídák’s multiple comparisons test. Correlation analysis was conducted using Spearman’s r-test. Simple Survival Analysis (Kaplan–Meier) was performed and Log-rank (Mantel-Cox) test was used for comparison between the two groups. The results are presented as means + standard error of mean (SEM) and p values < 0.05 were considered statistically significant.

## 3. Results

### 3.1. FIBCD1 protein is expressed in human CRC tissues and increases with disease progression

To determine whether FIBCD1 is expressed in human colorectal cancer tissue and to evaluate the extent of FIBCD1 expression in relation to disease progression, we performed immunohistochemical staining of colonic tumor specimens isolated from patients diagnosed with CRC stage I-III. In general, we found that FIBCD1 expression and distribution were highly variable across all three stages of disease severity assessed. However, direct assessment of the tissue specimens based on disease progression revealed a significant increase in FIBCD1 expression and distribution at more advanced tumor staging (p < 0.0398) (Figure 1A and B).

**Figure 1.**
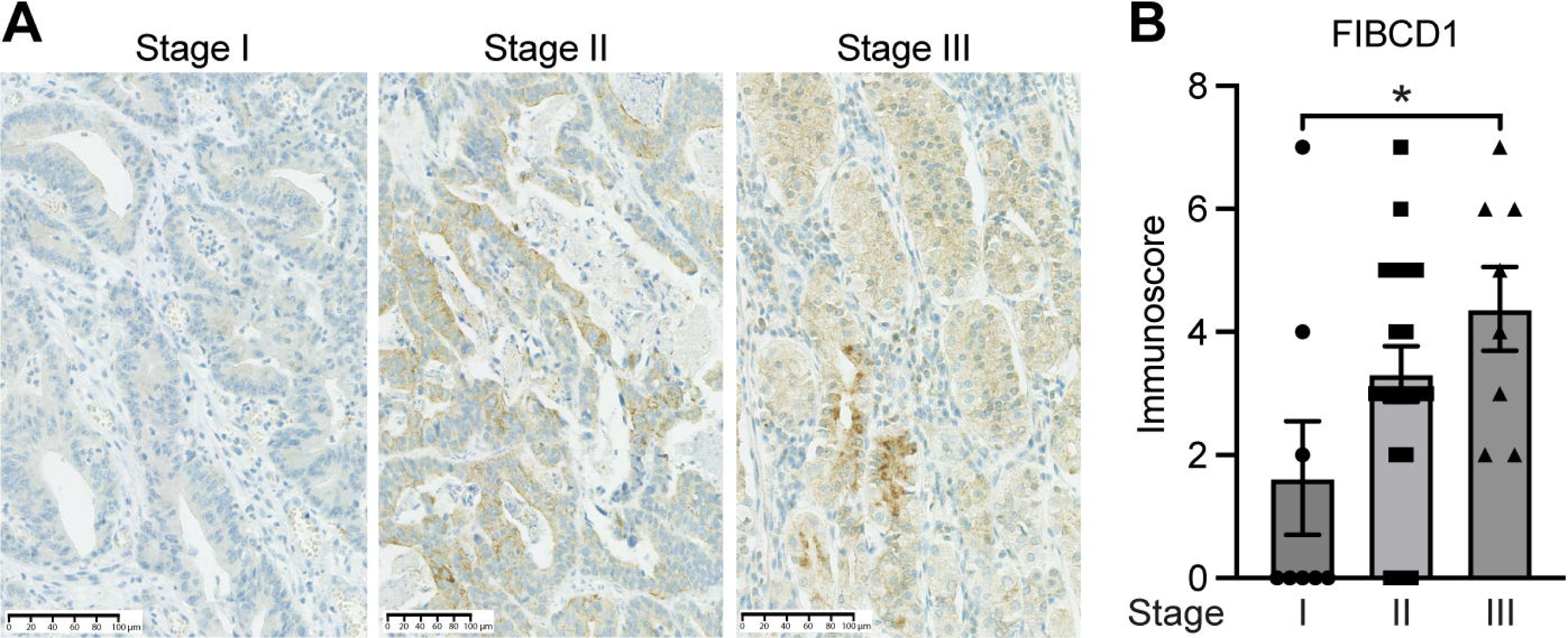
FIBCD1 expression in human colorectal cancer correlates with disease severity. **(A)** Representative immunohistochemical images of tumor tissues from patients with stage I-III CAC. Scalebar, 100 µm. **(B)** Statistical evaluation of FIBCD1 expression according to colorectal cancer stage I (n=8), II (n=19), and III (n=8). **Statistics:** Data are presented as mean ± SEM where dots represent individual specimens. One-way ANOVA followed by post hoc Tukey test for multiple comparisons. * P < 0.05.

### 3.2. Intestinal overexpression of FIBCD1 attenuates colonic inflammation in a mouse model for CAC

To investigate the potential significance of FIBCD1 on the development of CAC, we used *Fibcd1*-transgenic mice (overexpressing FIBCD1 in the intestinal epithelium) and WT littermates in an AOM/DSS mouse model of CAC (Figure 2A). A hallmark of experimental colitis is a notable body weight loss and shortening of the colon [22]. DSS-induced weight loss was significantly reduced following DSS treatments in TG mice compared to WT littermates (Figure 2B). In addition, Kaplan–Meier survival analysis revealed that TG mice had a higher survival rate compared to WT mice (p=0.006) (Figure 2C). At the time of the sacrifice, TG mice also showed significantly increased colon length compared to WT mice (p = 0.0007) (Figure 2D). Consistent with less severe disease symptoms, histopathological examination of the H&E-stained colon tissues revealed reduced intestinal colitis severity in TG mice, characterized by decreased infiltration of inflammatory cells and more normal-appearing overall mucosal architecture (p<0.0001) (Figure 2E).

**Figure 2.**
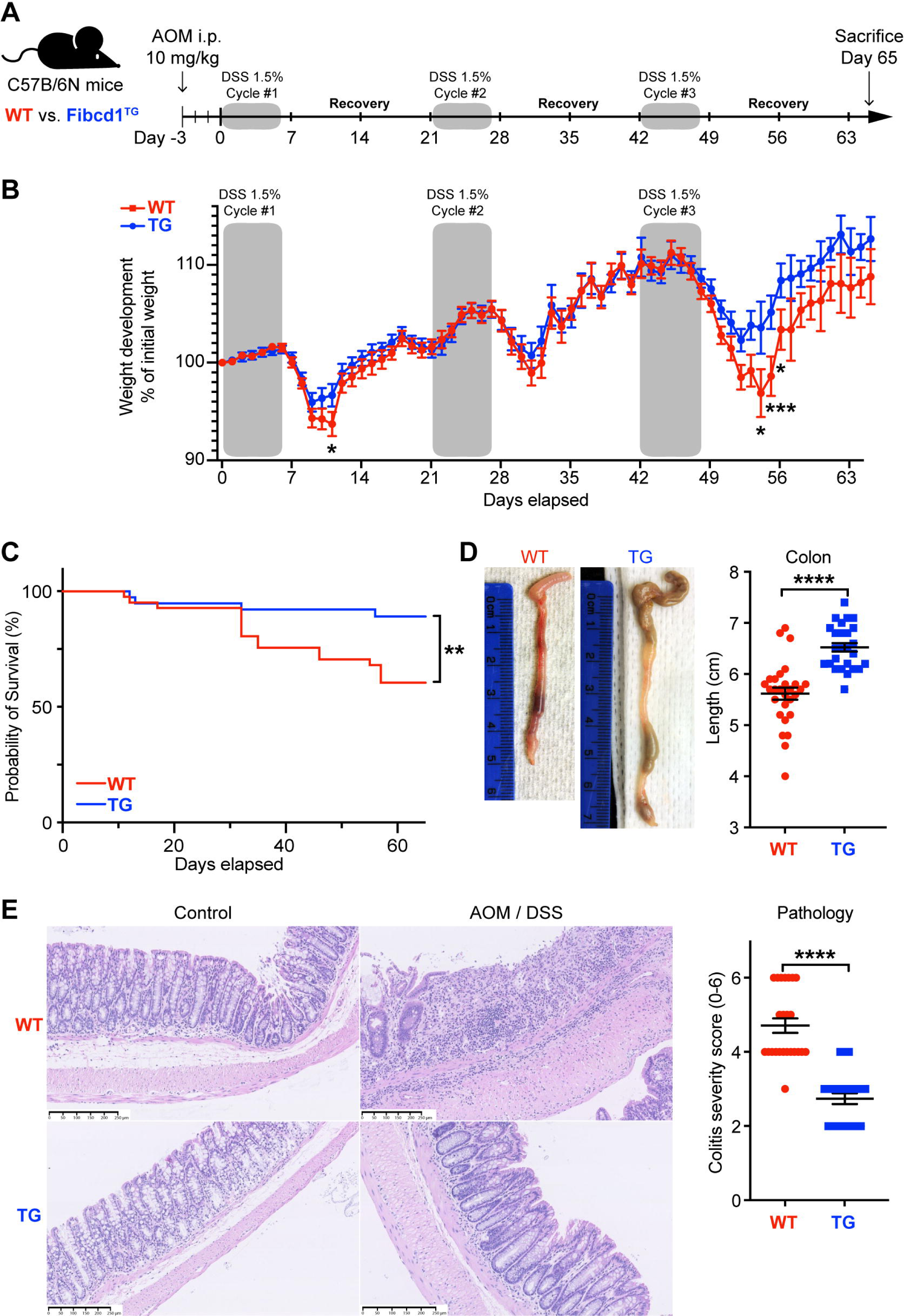
FIBCD1 overexpression attenuates colonic inflammation in a mouse model of colorectal cancer. **(A)** Schematic representation illustrating the experimental layout: Wildtype and *Fibcd1*-transgenic (TG) littermates received a single dose of AOM i.p. injection followed by three cycles of 1.5% DSS in drinking water. Mice were sacrificed 65 days after the experiment start. **(B)** Changes in the body weight of the AOM/DSS treated mice over the course of the experiment (n = 26-29 mice per group). **(C)** Kaplan-Meyer survival analysis of the WT and TG littermates exposed to AOM/DSS. **(D)** Representative images and statistics of colon lengths of WT and TG littermates at day 65 (n = 23-24 mice per group). Scalebar, 250 µm. **(E)** Representative H&E-stained images and evaluation of histological colitis severity (score for infiltration of inflammatory cells and colonic architecture) in colon tissue specimens from WT and TG littermates treated with AOM/DSS (n = 23-24 mice per group). Scale bar 250μm. **Statistics:** Results are pooled from three independent experiments and data are presented as mean ± SEM where dots represent individual mice. Mixed-effects model (REML) of Two-Way ANOVA repeated measures were performed for weight curves data in B, followed by post hoc Holm-Šídák’s multiple comparisons test. Simple Survival Analysis (Kaplan–Meier) was performed and the Log-rank (Mantel-Cox) test was used for the comparison of data in C. Unpaired Student’s t-test was performed to analyze data in D and E. * P < 0.05, ** P < 0.01, *** P < 0.001, and ****, P < 0.0001.

### 3.3. Overexpression of FIBCD1 ameliorates the progression of CAC

It is well-recognized that chronic inflammation of the intestinal mucosa can play a fundamental role in the initiation and progression of CRC [23]. To assess whether FIBCD1 expression influences tumor formation and development, the colons from WT and TG littermates sacrificed at day 65 in the CAC model were dissected longitudinally and stained with methylene blue. In these tissues, we observed a striking reduction in the number of visible tumors (p<0.0001) as well as tumor volume (p = 0.0019) in TG mice in comparison to WT mice (Figure 3A-C). Moreover, correlation analyses demonstrated a significant positive association between tumor load and histopathological colitis severity score (r = 0.679, p<0.0001), which was further supported by significant negative correlations between colon length vs. colitis severity (r = - 0.634, p<0.0001) as well as tumor load (r = - 0.505, p=0.0003) (Figure 3D to F).

**Figure 3.**
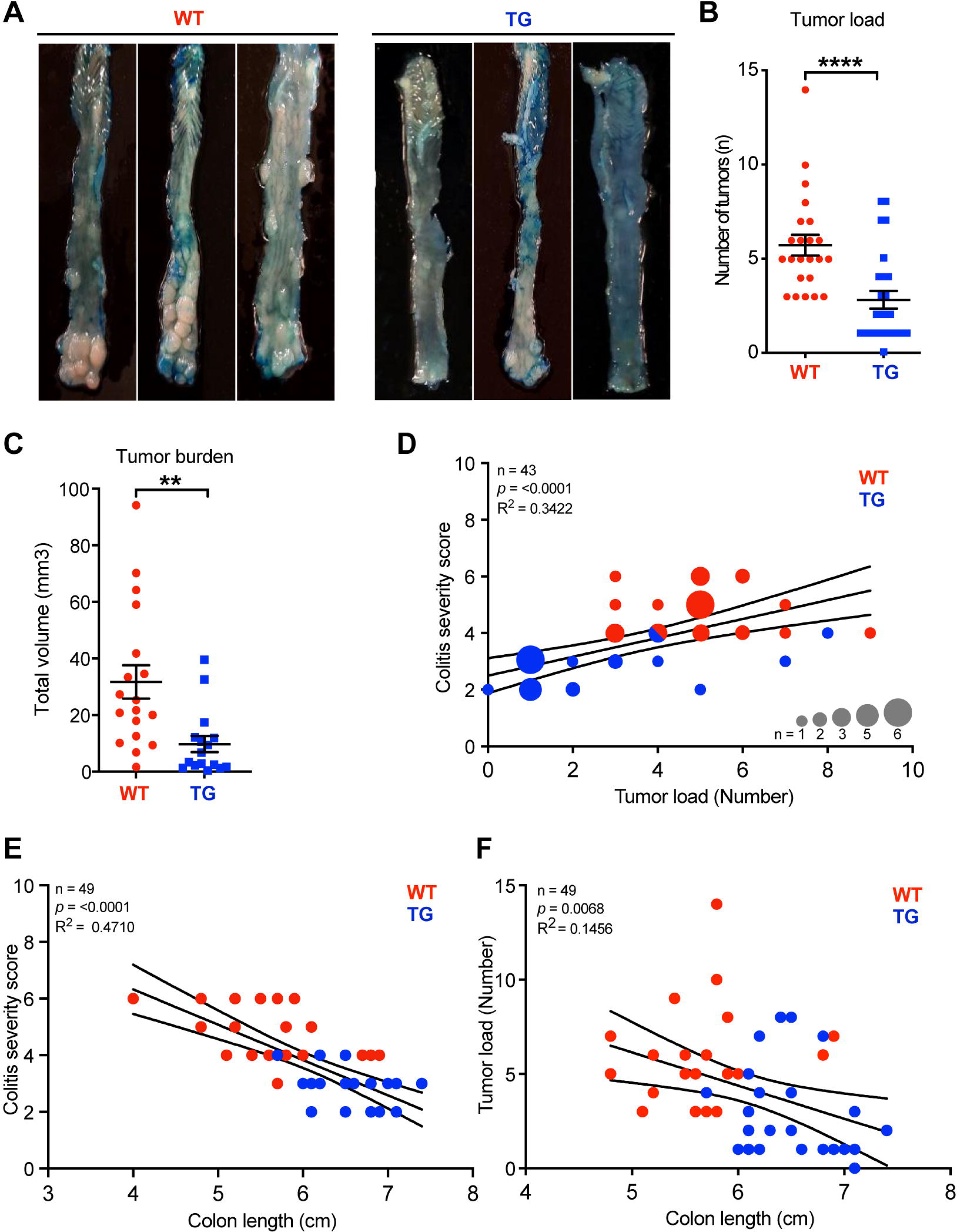
FIBCD1 overexpression ameliorates the development and progression of tumors in a mouse model of colorectal cancer. **(A)** Representative images of colonic tumors stained with methylene blue from WT and TG littermates at day 65. **(B and C)** Number and total volume of tumors in WT and TG littermates at the end of the experiment (n = 20-23 mice per group). **(D)** Correlation analysis between colitis severity score and tumor load, **(E)** between colitis severity score and colon length, and **(F)** between tumor load and colon length in WT and TG littermates (n = 43-49). **Statistics:** Results are pooled from two independent experiments and data are presented as mean ± SEM. The differences in dot size indicate the number of animals at that particular level. Unpaired Student’s t-test was performed to analyze data in B and C. nonparametric Spearman r test was performed to analyze data in D, E, and F. * P < 0.05, ** P < 0.01, *** P < 0.001, and ****, P < 0.0001.

### 3.4. FIBCD1 reduces the levels of biomarkers associated with intestinal disease and inflammation

Fecal biomarkers such as calprotectin and lipocalin-2 have been identified as important diagnostic tools to evaluate inflammatory activity in the intestinal mucosa [24]. To further examine the impact of FIBCD1 on disease severity, fecal samples were collected from WT and TG mice over the course of the experiment. We found that TG mice had significantly lower levels of LCN2 and CAL in the feces compared to WT littermates at several time points (Figure 4A and B). To examine the impact of FIBCD1 at the transcriptional level, RNA was isolated from the dissected colonic tumors and the tumor-adjacent tissue for RT-qPCR analysis. In the tumor tissues, we observed reduced expression of inflammatory genes in the TG mice in comparison to WT littermates. Expression levels of *Il1b*, one of the major mediators of inflammatory responses, as well as the pleiotropic cytokine *Il6,* were both significantly reduced in the tumors of TG mice compared to WT mice. Likewise, mRNA levels for inflammatory markers such as *Il17*, *Tnfa,* and *Ccl2 as* well as the anti-microbial peptide *Reg3g* were significantly downregulated in TG mice (Figure 4C). We also investigated the gene expression profile in the tumor-adjacent tissue and observed a similar response profile, characterized by a general decrease in pro-inflammatory signature, with downregulation of transcripts for *Il1b, Il6, Il17, Ccl2*, as well as reduced expression of *Il2* in the TG mice (Supplementary Figure 1).

**Figure 4.**
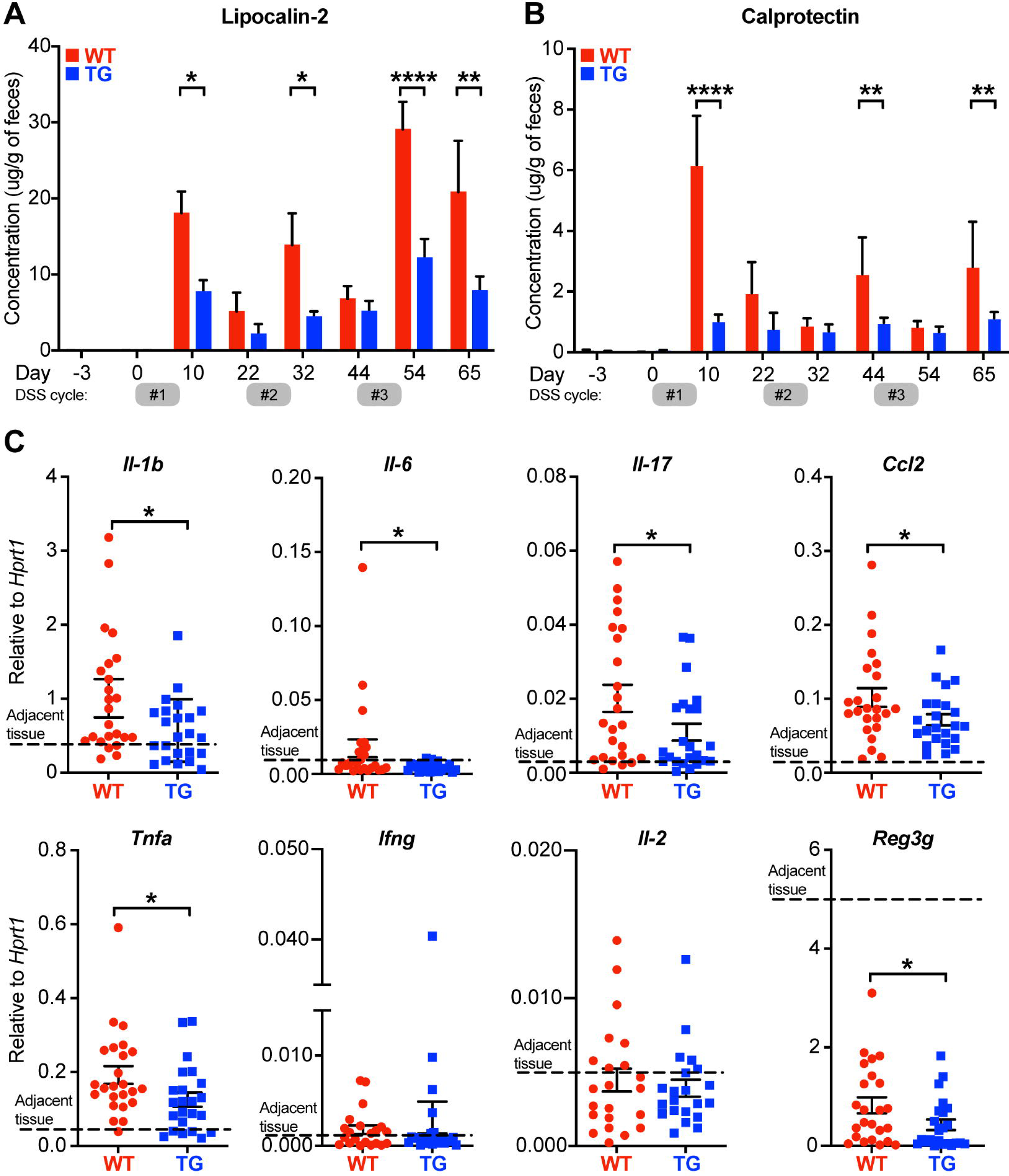
FIBCD1 influences biomarkers associated with disease and inflammation. **(A, B)** LCN2 and Calprotectin (inflammatory biomarkers) were quantified in fecal specimens before disease induction and at different time points throughout the experiment (n = 12 samples per group). **(C)** Relative expression of the indicated genes in tumor tissues isolated from WT and TG littermates (n=23-24 mice per group). The dotted line represents the average level of expression in adjacent tissues (WT and TG combined). **Statistics:** Results are pooled from two independent experiments and data are presented as mean ± SEM where dots represent individual mice. Mixed-effects model (REML) of Two-Way ANOVA repeated measures were performed for data in A and B, followed by post hoc Holm-Šídák’s multiple comparisons test. Unpaired Student’s t-test was performed to analyze data in C. * p < 0.05, ** p < 0.01, and ****, p < 0.0001.

### 3.5. FIBCD1 alters the expression of apoptosis-related genes in colonic tumors

To further assess the effects of FIBCD1 on CAC tumor progression, we analyzed the isolated tumor tissues from AOM/DSS-treated WT and TG littermates for the expression of CAC-related oncogenes and tumor suppressor genes as well as genes involved in cell proliferation and apoptosis. When quantified we found no significant difference in the expression of well-known oncogenes like *Kras*, *Erbb2, Sox9, Ctnnb1, Stat3*, and *Pik3ca* between the tumor tissues isolated from WT and TG littermates (Figure 5A). Moreover, we found no difference in the expression of genes related to tumor suppression like *Apc, Tp53, Tgfbr2,* and *Smad4* (Figure 5B). In contrast, genes associated with apoptosis like *Bcl2*, *Xiap*, and *Casp1* were found to be marginally yet significantly upregulated in TG when compared to WT littermates (Figure 5C). Additionally, the expression of these cancer-related genes was also assessed in adjacent tissues, where no changes between WT and TG littermates were observed (Supplementary Figure 2).

**Figure 5.**
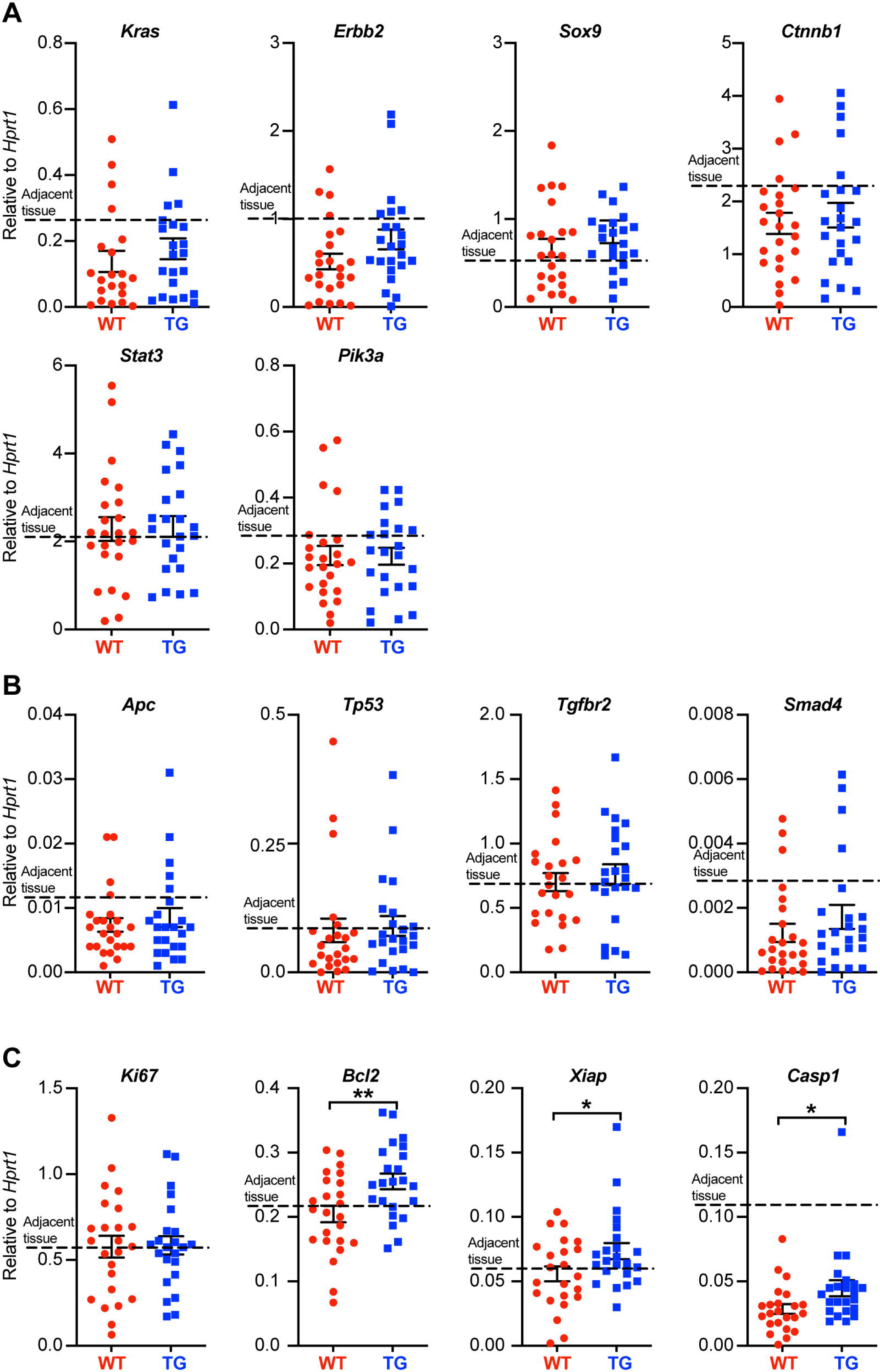
FIBCD1 alters the expression of genes associated with apoptosis in colonic tumor tissues. **(A)** Relative expression of the indicated oncogenes, **(B)** tumor suppressor genes, and **(C)** genes associated with cell proliferation and apoptosis in tumor tissues isolated from WT and TG littermates (n=23-24 mice per group). The dotted line represents the average level of expression in adjacent tissues (WT and TG combined). **Statistics:** Results are pooled from two independent experiments and data are presented as mean ± SEM where dots represent individual mice. Unpaired Student’s t test was performed to analyze data. * P < 0.05 and ** P < 0.01.

### 3.6. FIBCD1’s protective effect against CAC is not mediated by compositional changes in the intestinal microbiota

FIBCD1 is a chitin-binding receptor that has previously been shown to limit and control fungal dysbiosis in the intestines [14]. Following, we asked whether the protective effect of FIBCD1 on the development of CAC could be partially mediated by compositional changes to the intestinal microbiota. To investigate this, we purified fecal DNA from WT and TG mice and evaluated changes in the microbiota by qPCR analysis. In general, we observed a very limited presence of fungal DNA in the collected fecal samples (fungal Ct values were consistently above 32 or non-detected) in all sample material assessed using well-established quantitative procedures [14, 25]. Moreover, we found no significant differences in the overall fungal burden between co-housed TG and WT littermates regardless of disease progression (Figure 6A). Similarly, we did not detect significant changes in the abundance of the biggest bacterial phyla including *Firmicutes* and *Bacteroides* at any time point (Figure 6B and C).

**Figure 6.**
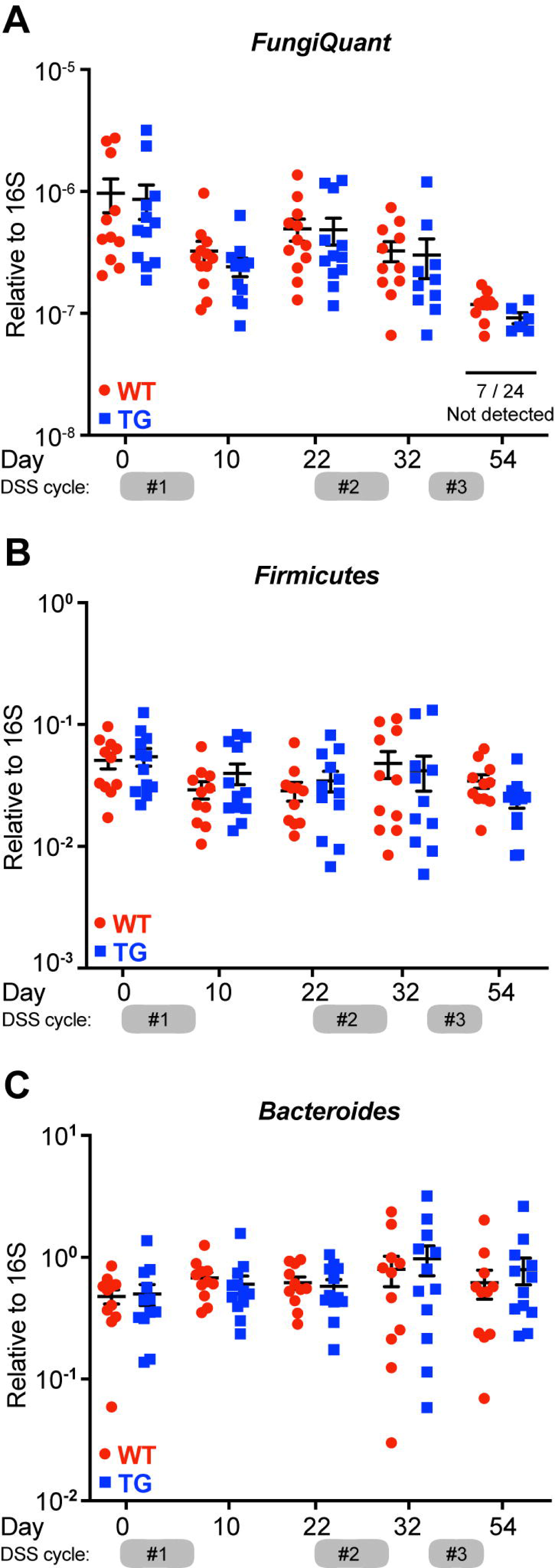
FIBCD1 protective effects are independent of the intestinal microbiota composition. **(A)** Quantification of total fungal presence in feces from co-housed WT and TG littermates using broad coverage primers (FungiQuant) at different time points throughout the experiment (n = 12 samples per group). **(B and C)** Quantification of relative abundance of *Firmicutes* and *Bacteroides* relative to total 16S in feces from co-housed WT and TG littermates at different time points throughout the experiment (n = 12 samples per group). **Statistics:** Results are pooled from two independent experiments and data are presented as mean ± SEM where dots represent individual mice. No significant differences were observed when data were analyzed using a mixed-effects model (REML) of Two-Way ANOVA repeated measures followed by post hoc Holm-Šídák’s multiple comparisons test.

### 3.7. Soluble FIBCD1 is detected in feces from transgenic mice and increases during acute inflammation

Recently, Graca et al. have identified FIBCD1 as a possible myokine that is shed from the surface of cells expressing full-length transmembrane FIBCD1 [26]. To investigate FIBCD1 as a potential shedded marker of inflammation in the context of CRC, we developed a sandwich ELISA that recognizes epitopes contained in a shedded variant of FIBCD1 (Figure 7A). After validation and optimization of the assay, we assessed for the presence of shedded FIBCD1 in fecal sample supernatants collected from WT and TG littermates in the AOM/DSS mouse model of CAC. In accordance with previous reports of very low expression of *Fibcd1* in mouse intestinal tissues [14], our assay did not detect any presence of shedded FIBCD1 protein in the sample material collected from WT mice. However, in sample material collected from TG littermates, we observed a high concentration of shedded FIBCD1 across all samples evaluated (Figure 7B). Moreover, we observed a highly significant increase in shedded FIBCD1 in samples collected from TG littermates during the acute phase (day 10) of the initial inflammation induced by DSS treatment (p<0.0001).

**Figure 7.**
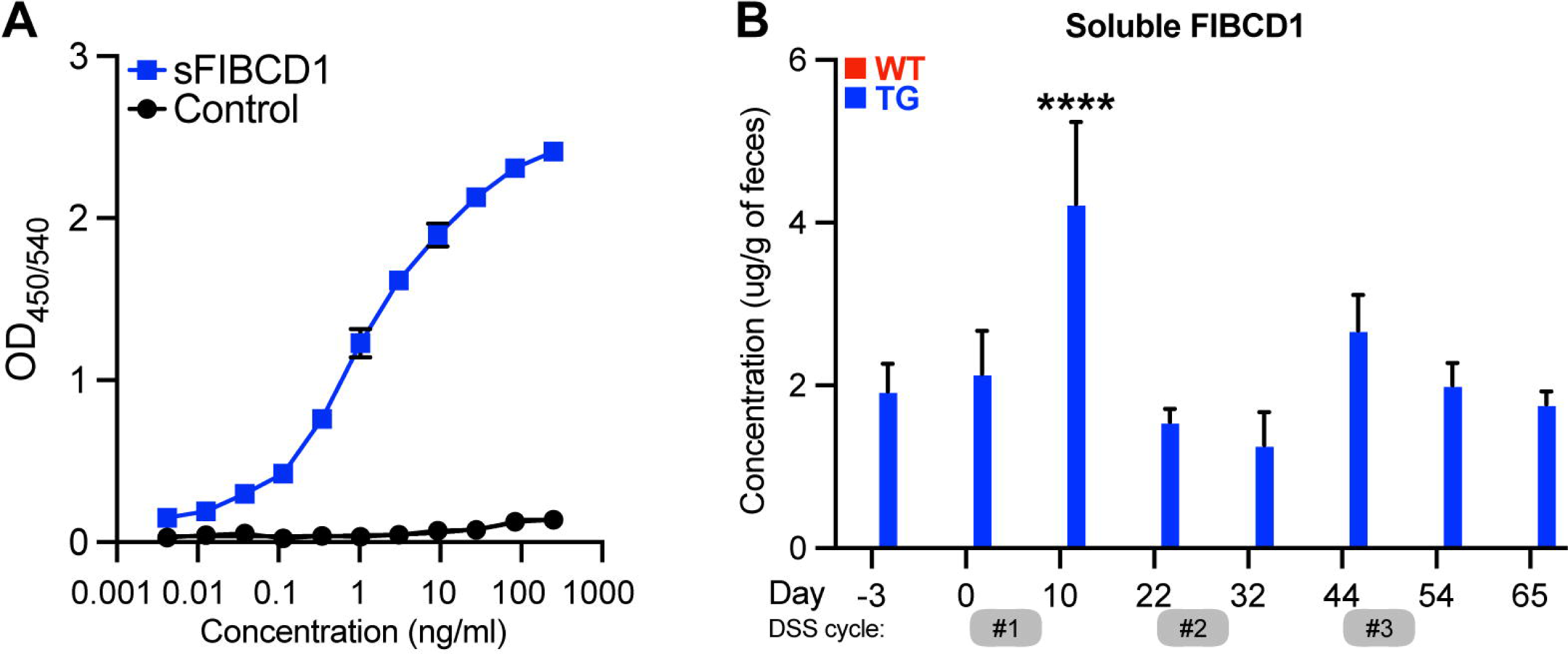
Soluble FIBCD1 is detectable in feces at increased levels during acute inflammation. **(A)** Representative binding curves demonstrating the ELISA developed for detection and quantification of soluble FIBCD1. Recombinant FIBCD1 was used as a standard (0-250 ng) and resuspended in a fixed amount of fecal sample suspension (10mg/ml). **(B)** Quantification of soluble FIBCD1 in feces isolated from WT and TG littermates at different time points throughout the experiment (n = 12 samples per group). **Statistics:** Results are pooled from two independent experiments and data are presented as mean ± SEM. All samples from WT mice were below detection. Mixed-effects model (REML) of Two-Way ANOVA repeated measures were performed for TG sample groups, followed by post hoc Holm-Šídák’s multiple comparisons test. ****, P < 0.0001.

## 4. Discussion

In the present study, we sought to examine the role of intestinal FIBCD1 in CRC. In human material from CRC patients, we identified expression of FIBCD1 in the tumor tissues where expression and distribution were found to correlate with the disease stage, supporting a possible role for FIBCD1 in human CRC. Utilizing transgenic mice overexpressing FIBCD1 in the intestinal tissues, thereby mimicking its expression in the human gut, we found that expression of FIBCD1 significantly alleviated intestinal inflammation, increased survival, and decreased tumor formation as well as burden in a mouse model of inflammation-driven colorectal cancer. Moreover, we observed that the reduction of disease severity was associated with decreased expression of several canonical inflammatory mediators implicated in the development and progression of CAC [27, 28]. On the other hand, overexpression of FIBCD1 did not exhibit a notable impact on the expression of genes associated with tumor development or suppression. Additionally, FIBCD1 did not appear to mediate disease protection by inducing significant fungal or bacterial alterations in the gut microbiota. Interestingly, we observed the presence of soluble FIBCD1 protein in the feces of TG mice, with a significant increase in the initial phases of intestinal inflammation. These findings suggest the potential use of shedded FIBCD1 as a biomarker of inflammation and/or disease progression in future applications.

By employing the AOM/DSS mouse model, we present novel evidence that increased expression of FIBCD1 in the intestines significantly attenuated the development of CAC. This attenuation was characterized by a decrease in the number of tumors and overall tumor burden. Chronic inflammation often precedes cancer development and plays a decisive role in various steps involved in cancer initiation and progression [29, 30]. It is well established that aberrant inflammatory responses in the tumor microenvironment can promote cancer cell survival, neoangiogenesis, and metastatic spread as well as interfere with anti-cancer therapy [29]. FIBCD1 overexpression in IECs resulted in attenuated intestinal inflammation characterized by reduced weight loss, increased colon length, and decreased epithelial damage. Furthermore, we observed reduced levels of calprotectin and lipocalin-2 in FIBCD1-overexpressing mice. Both calprotectin and lipocalin-2 are important fecal biomarkers associated with intestinal disease and inflammation [24]. Previous studies have shown that fecal lipocalin-2 is elevated in patients with CRC and associated with tumor progression [31, 32]. Similarly, high levels of fecal calprotectin have been shown to be a pre-diagnostic factor for patients with CRC and positively correlate with tumor growth [33].

The observed reduction in tumor burden and tumor size in TG mice was associated with decreased expression of inflammatory cytokines such as *Il-1b*, *Il-6*, *Il-17*, and *Ccl2* in the tumor and tumor-adjacent tissue. Inflammatory mediators in the tumor microenvironment can enhance tumor development via the recruitment of additional immune cells that in turn provide survival signals and growth factors for cancer cells [4, 29]. For example, IL-6 expression has been shown to drive early tumorigenesis in CAC via activation of the STAT3 pathway that increases the survival of pre-malignant IECs [34]. IL-17 has been shown to have a pro-tumorigenic effect in CRC development as well as promote resistance to anti-cancer therapy [35]. As inflammation is instrumental in tumor development in CAC, the ability of FIBCD1 to modulate and dampen inflammatory responses in the intestinal tract may be one of its primary anti-tumorigenic actions observed in the AOM/DSS model. On the other hand, high levels of FIBCD1 expression in tissue samples from patients with gastric and hepatic cancer have been associated with increased tumor size and invasiveness as well as overall poorer prognosis and survival [36, 37]. In agreement with this, our analysis of colorectal cancer tissue showed increased expression of FIBCD1 at more advanced tumor stages. A possible explanation for this discrepancy may be that during tumor progression, as a means of immune escape mechanism, carcinogenic epithelial cells increase expression of FIBCD1 to induce a pro-tumor microenvironment characterized by increased local immune suppression that favors cancer growth and metastasis [3]. Hence, FIBCD1 might be an emerging novel molecular target and/or a prognostic biomarker of several different gastrointestinal malignancies.

The anti-inflammatory properties of FIBCD1 reported in previous studies have been attributed to direct fungal recognition and reduced fungal outgrowth [14, 16, 38]. Additionally, mycobiota sequence analyses showed compositional changes in the fungal microbiota in FIBCD1-overexpressing mice compared to WT littermates [14]. Emerging research has shown an association between fungal dysbiosis and CRC development and progression [39-41]. Increased burden of *Candida* spp. as well as altered composition of various fungal communities have been identified in CRC patients compared to healthy controls [42]. In this study, we generally observed a very limited fungal presence in co-housed WT and TG littermates maintained in our animal facility. Thus, we were not able to detect significant changes in the total abundance of fungal DNA or prevalent species like *Candida* spp., suggesting that the reduced intestinal inflammation observed in TG mice was not linked to decreased fungal burden or changes in fungal composition observed in previous studies [14].

Nevertheless, the anti-tumor properties of FIBCD1 were evident in the AOM/DSS model, suggesting an alternative mechanism for the protective effect of FIBCD1 in inflammation-associated carcinogenesis. In our study, we found no evidence of FIBCD1 expression directly influencing the expression of well-known oncogenes or tumor suppressor genes within the tumor tissues. On the other hand, we observed a slight but significant difference in apoptosis-related gene expression, which could be contributed to various reasons including changes in the tumor-associated immune cell composition. However, Graca et al. have recently demonstrated that FIBCD1 can be cleaved from the cell surface, generating a secreted form of FIBCD1 protein containing the fibrinogen-related domain that participates in ligand binding [26]. Soluble FIBCD1 has been shown to interact with and regulate integrin subunits as well as reduce expression of inflammatory genes in skeletal muscle cells during cancer-induced myofiber atrophy [26]. Building upon these findings, we developed a sandwich ELISA to evaluate the potential presence of FIBCD1 in feces due to the shedding of epithelial-derived membrane-bound FIBCD1 in response to inflammation and the development of CAC. While we were not able to detect and quantify endogenous soluble FIBCD1 in feces from WT mice, our assay proved sensitive for the detection and quantification of soluble FIBCD1 in fecal specimens from TG mice. Proteolytic cleavage of membrane proteins is a fundamental molecular process that serves as a functional switch of hundreds of target proteins involved in most physiological processes as well as cancer and inflammation [43, 44]. In our animal model, we observed a highly significant increase of soluble FIBCD1 in feces during the initial phase of inflammation and disease development. Although transgenic, and thereby highly artificial by nature, the discovery of a soluble variant of FIBCD1 derived from membrane-bound protein may be highly relevant for the understanding of FIBCD1 function in relation to human inflammation and CAC development. Moreover, the data provoke the question of whether soluble FIBCD1 might serve as a novel noninvasive biomarker of disease development in human pathologies like IBD or CAC.

In a recent study by Fell et al., FIBCD1 was shown to function as an endocytic receptor for glycosaminoglycans, thereby regulating signaling within the brain’s extracellular matrix and playing a pivotal role in the structure and function of the nervous system [45]. Notably, mice lacking *Fibcd1* exhibited impaired hippocampal-dependent learning and diminished synaptic remodeling, underscoring the significant involvement of FIBCD1 in shaping the structural and functional aspects of the nervous system. Furthermore, single-cell and spatial transcriptomic sequencing of hippocampal tissue from mice with Parkinson’s disease identified *Fibcd1* as one of the highly differentially regulated genes in the CA3 subregion, involved in memory and cognitive functions [46, 47]. These studies further illustrate that FIBCD1 exhibits pleiotropic functions, with emerging novel roles in various diseases and physiological functions extending beyond chitin recognition.

Taken together, the present study using an AOM/DSS-induced CAC mouse model found that overexpression of FIBCD1 in the intestinal epithelium resulted in decreased inflammation and tumor formation. The presence of soluble FIBCD1 during the initial stages of intestinal inflammation introduces intriguing possibilities concerning its potential as an immune response regulator during inflammation. It is conceivable that shedded FIBCD1 contributes to the intricate regulation of the inflammatory milieu by facilitating immune cell recruitment and migration through integrin binding or by selectively binding to specific cell types and influencing their transcriptomic profile, thus mitigating inflammatory responses. Understanding the precise role and mechanisms of action of soluble FIBCD1 has potential to pave the way for innovative therapeutic approaches targeting inflammation-related diseases. Given the well-established connection between inflammation and cancer development, it is plausible that FIBCD1 may hold anti-carcinogenic properties that stem from its ability to influence early inflammatory responses in the intestinal tract. Although the mechanistic basis underlying this protective effect remains to be determined, our observations suggest that FIBCD1 might be a novel molecular target in CAC and an attractive candidate for future research.

## Data Availability

All data produced in the present study are available upon reasonable request to the authors

## Acknowledgments

We thank HSSM lab members for discussion and critical reading of the manuscript.

## Author contributions

V.K.S., M.D., B.P, S.S., S.G.S., S.D., G.L.S., U.H., and J.B.M. designed and performed the research. V.K.S., M.D., S.D., and J.B.M. analyzed the data. B.B. collected and provided the human CRC samples. V.K.S., M.D., and J.B.M. wrote the manuscript with input from the other authors.

## Figure legends

**Supplementary Figure 1. FIBCD1 ameliorates the expression of inflammatory markers in non-tumorigenic adjacent tissues.** Relative expression of the indicated genes in non-tumorigenic adjacent tissues isolated from WT and TG littermates (n=23-24 mice per group). The dotted line indicates the average level of expression in non-treated colonic tissues isolated from naïve WT and TG littermate controls. **Statistics:** Results are pooled from two independent experiments and data are presented as mean ± SEM where dots represent individual mice. Unpaired Student’s t test was performed to analyze data. * p < 0.05.

**Supplementary Figure 2. FIBCD1 expression does not influence genes related to CAC in non-tumorigenic adjacent tissues. (A)** Relative expression of the indicated oncogenes, **(B)** tumor suppressor genes, and **(C)** genes associated with cell proliferation and apoptosis in non-tumorigenic adjacent tissues isolated from WT and TG littermates (n=23-24 mice per group). The dotted line indicates the average level of expression in non-treated colonic tissues isolated from naïve WT and TG littermate controls. **Statistics:** Results are pooled from two independent experiments and data are presented as mean ± SEM where dots represent individual mice. No significant differences were observed when Unpaired Student’s t-test was performed to analyze data in (A, B, and C).

**Table 1.** TaqMan primers.

| Gene symbol | Description | Catalog number |
| --- | --- | --- |
| HPRT1 | Hypoxanthine-guanine phosphoribosyltransferase | Mm00446968_m1 |
| IL-1b | Interleukin-1 beta | Mm00434228 |
| IL-6 | Interleukin-6 | Mm00446190 |
| Ifng | Interferon gamma | Mm00801778_m1 |
| Reg3g | Regenerating islet-derived protein 3 gamma | Mm00441127_m1 |
| Tgfb2 | Transforming growth factor-beta 2 | Mm00436955_m1 |
| Bcl2 | B-cell lymphoma 2 | Mm00477631_m1 |
| Xiap | X-linked inhibitor of apoptosis protein | Mm01311594_m1 |
| Casp1 | Caspase 1 | Mm00438023_m1 |

**Table 2.**
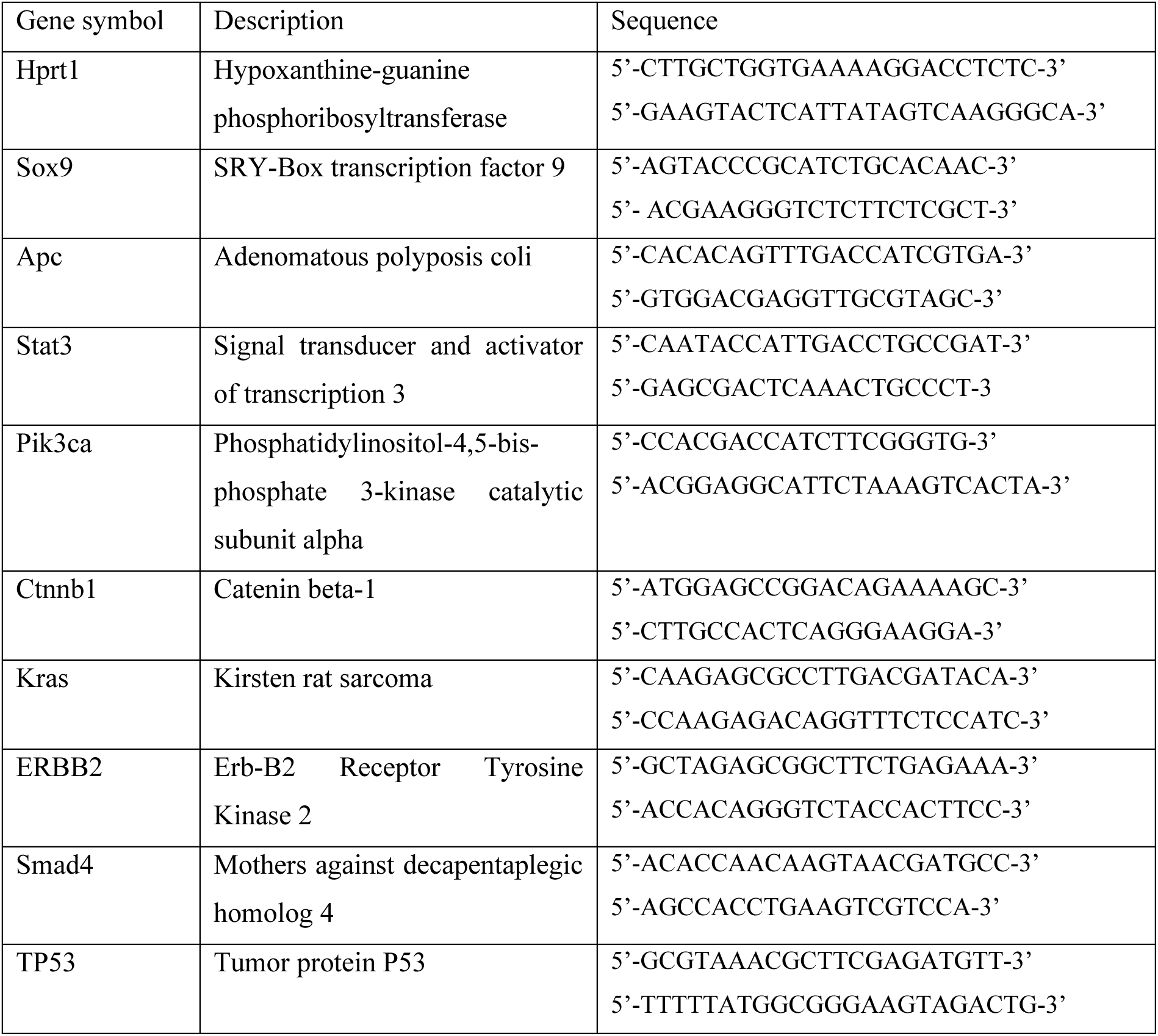
SYBR-green primers.

## References

1. Rawla, P., T. Sunkara, and A. Barsouk, Epidemiology of colorectal cancer: incidence, mortality, survival, and risk factors. Prz Gastroenterol, 2019. 14(2): p. 89–103.

2. Mantovani, A., et al., Cancer-related inflammation. Nature, 2008. 454(7203): p. 436–44.

3. Zhao, H., et al., Inflammation and tumor progression: signaling pathways and targeted intervention. Signal Transduct Target Ther, 2021. 6(1): p. 263.

4. Greten, F.R. and S.I. Grivennikov, Inflammation and Cancer: Triggers, Mechanisms, and Consequences. Immunity, 2019. 51(1): p. 27–41.

5. Porter, R.J., et al., Inflammatory Bowel Disease-Associated Colorectal Cancer: Translational Risks from Mechanisms to Medicines. J Crohns Colitis, 2021. 15(12): p. 2131–2141.

6. Kruk, J. and H.Y. Aboul-Enein, Reactive Oxygen and Nitrogen Species in Carcinogenesis: Implications of Oxidative Stress on the Progression and Development of Several Cancer Types. Mini Rev Med Chem, 2017. 17(11): p. 904–919.

7. Vetter, L.E., et al., Colorectal cancer in Crohn’s colitis is associated with advanced tumor invasion and a poorer survival compared with ulcerative colitis: a retrospective dual-center study. Int J Colorectal Dis, 2021. 36(1): p. 141–150.

8. Soh, J.S., et al., Immunoprofiling of Colitis-associated and Sporadic Colorectal Cancer and its Clinical Significance. Sci Rep, 2019. 9(1): p. 6833.

9. Parang, B., C.W. Barrett, and C.S. Williams, AOM/DSS Model of Colitis-Associated Cancer. Methods Mol Biol, 2016. 1422: p. 297–307.

10. De Robertis, M., et al., The AOM/DSS murine model for the study of colon carcinogenesis: From pathways to diagnosis and therapy studies. J Carcinog, 2011. 10: p. 9.

11. Thaker, A.I., et al., Modeling colitis-associated cancer with azoxymethane (AOM) and dextran sulfate sodium (DSS). J Vis Exp, 2012(67).

12. von Huth, S., et al., Immunohistochemical Localization of Fibrinogen C Domain Containing 1 on Epithelial and Mucosal Surfaces in Human Tissues. J Histochem Cytochem, 2018. 66(2): p. 85–97.

13. Schlosser, A., et al., Characterization of FIBCD1 as an acetyl group-binding receptor that binds chitin. J Immunol, 2009. 183(6): p. 3800–9.

14. Moeller, J.B., et al., Modulation of the fungal mycobiome is regulated by the chitin-binding receptor FIBCD1. J Exp Med, 2019. 216(12): p. 2689–2700.

15. Andersen, M.C.E., et al., FIBCD1 ameliorates weight loss in chemotherapy-induced murine mucositis. Support Care Cancer, 2021. 29(5): p. 2415–2421.

16. Jepsen, C.S., et al., FIBCD1 Binds Aspergillus fumigatus and Regulates Lung Epithelial Response to Cell Wall Components. Front Immunol, 2018. 9: p. 1967.

17. Bhattacharya, S., et al., FIBCD1 Deficiency Decreases Disease Severity in a Murine Model of Invasive Pulmonary Aspergillosis. Immunohorizons, 2021. 5(12): p. 983–993.

18. Viennois, E., et al., Dextran sodium sulfate inhibits the activities of both polymerase and reverse transcriptase: lithium chloride purification, a rapid and efficient technique to purify RNA. BMC Res Notes, 2013. 6: p. 360.

19. Krych, Ł., et al., Have you tried spermine? A rapid and cost-effective method to eliminate dextran sodium sulfate inhibition of PCR and RT-PCR. J Microbiol Methods, 2018. 144: p. 1–7.

20. Erben, U., et al., A guide to histomorphological evaluation of intestinal inflammation in mouse models. Int J Clin Exp Pathol, 2014. 7(8): p. 4557–76.

21. Nexoe, A.B., et al., Colonic Epithelial Surfactant Protein D Expression Correlates with Inflammation in Clinical Colonic Inflammatory Bowel Disease. Inflamm Bowel Dis, 2019. 25(8): p. 1349–1356.

22. Chassaing, B., et al., Dextran sulfate sodium (DSS)-induced colitis in mice. Curr Protoc Immunol, 2014. 104: p. 15.25.1–15.25.14.

23. Schmitt, M. and F.R. Greten, The inflammatory pathogenesis of colorectal cancer. Nat Rev Immunol, 2021. 21(10): p. 653–667.

24. Zollner, A., et al., Faecal Biomarkers in Inflammatory Bowel Diseases: Calprotectin Versus Lipocalin-2-a Comparative Study. J Crohns Colitis, 2021. 15(1): p. 43–54.

25. Liu, C.M., et al., FungiQuant: a broad-coverage fungal quantitative real-time PCR assay. BMC Microbiol, 2012. 12: p. 255.

26. Graca, F.A., et al., The myokine Fibcd1 is an endogenous determinant of myofiber size and mitigates cancer-induced myofiber atrophy. Nat Commun, 2022. 13(1): p. 2370.

27. Knüpfer, H. and R. Preiss, Serum interleukin-6 levels in colorectal cancer patients--a summary of published results. Int J Colorectal Dis, 2010. 25(2): p. 135–40.

28. Francescone, R., V. Hou, and S.I. Grivennikov, Cytokines, IBD, and colitis-associated cancer. Inflamm Bowel Dis, 2015. 21(2): p. 409–18.

29. Grivennikov, S.I., F.R. Greten, and M. Karin, Immunity, inflammation, and cancer. Cell, 2010. 140(6): p. 883–99.

30. Afify, S.M., et al., Cancer-inducing niche: the force of chronic inflammation. Br J Cancer, 2022. 127(2): p. 193–201.

31. McLean, M.H., et al., Expression of neutrophil gelatinase-associated lipocalin in colorectal neoplastic progression: a marker of malignant potential? Br J Cancer, 2013. 108(12): p. 2537–41.

32. Chaudhary, N., et al., Lipocalin 2 expression promotes tumor progression and therapy resistance by inhibiting ferroptosis in colorectal cancer. Int J Cancer, 2021. 149(7): p. 1495–1511.

33. Blad, N., R. Palmqvist, and P. Karling, Pre-diagnostic faecal calprotectin levels in patients with colorectal cancer: a retrospective study. BMC Cancer, 2022. 22(1): p. 315.

34. Grivennikov, S., et al., IL-6 and Stat3 are required for survival of intestinal epithelial cells and development of colitis-associated cancer. Cancer Cell, 2009. 15(2): p. 103–13.

35. Wang, K., et al., Interleukin-17 receptor a signaling in transformed enterocytes promotes early colorectal tumorigenesis. Immunity, 2014. 41(6): p. 1052–63.

36. Jiang, C., et al., Overexpression of FIBCD1 Is Predictive of Poor Prognosis in Gastric Cancer. Am J Clin Pathol, 2018. 149(6): p. 474–483.

37. Wang, Y., et al., FIBCD1 overexpression predicts poor prognosis in patients with hepatocellular carcinoma. Oncol Lett, 2020. 19(1): p. 795–804.

38. Pilecki, B. and J.B. Moeller, Fungal recognition by mammalian fibrinogen-related proteins. Scand J Immunol, 2020. 92(4): p. e12925.

39. Wang, Y., et al., Fungal dysbiosis of the gut microbiota is associated with colorectal cancer in Chinese patients. Am J Transl Res, 2021. 13(10): p. 11287–11301.

40. Zhang, Z., et al., Gut fungi enhances immunosuppressive function of myeloid-derived suppressor cells by activating PKM2-dependent glycolysis to promote colorectal tumorigenesis. Exp Hematol Oncol, 2022. 11(1): p. 88.

41. Luan, C., et al., Dysbiosis of fungal microbiota in the intestinal mucosa of patients with colorectal adenomas. Sci Rep, 2015. 5: p. 7980.

42. Coker, O.O., et al., Enteric fungal microbiota dysbiosis and ecological alterations in colorectal cancer. Gut, 2019. 68(4): p. 654–662.

43. Lichtenthaler, S.F., M.K. Lemberg, and R. Fluhrer, Proteolytic ectodomain shedding of membrane proteins in mammals-hardware, concepts, and recent developments. Embo j, 2018. 37(15).

44. Düsterhöft, S., et al., Membrane-proximal domain of a disintegrin and metalloprotease-17 represents the putative molecular switch of its shedding activity operated by protein-disulfide isomerase. J Am Chem Soc, 2013. 135(15): p. 5776–81.

45. Fell, C.W., et al., FIBCD1 is an endocytic GAG receptor associated with a novel neurodevelopmental disorder. EMBO Mol Med, 2022. 14(9): p. e15829.

46. Jia, E., et al., Spatial Transcriptome Profiling of Mouse Hippocampal Single Cell Microzone in Parkinson’s Disease. Int J Mol Sci, 2023. 24(3).

47. Kesner, R.P., Behavioral functions of the CA3 subregion of the hippocampus. Learn Mem, 2007. 14(11): p. 771–81.

