## Supplementary figures and images for "Expression of FIBCD1 by intestinal epithelial cells alleviates inflammation-driven tumorigenesis in a mouse model of colorectal cancer"

### Supplemental figures

#### Supplementary figure 1

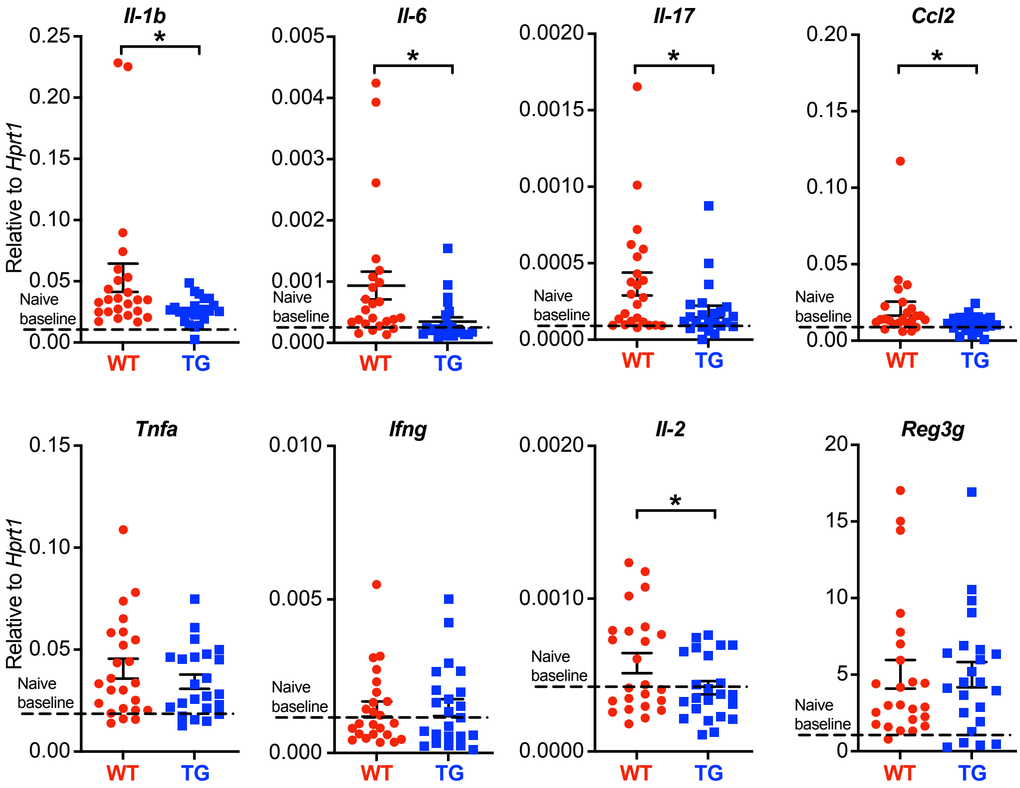

#### Supplementary figure 2

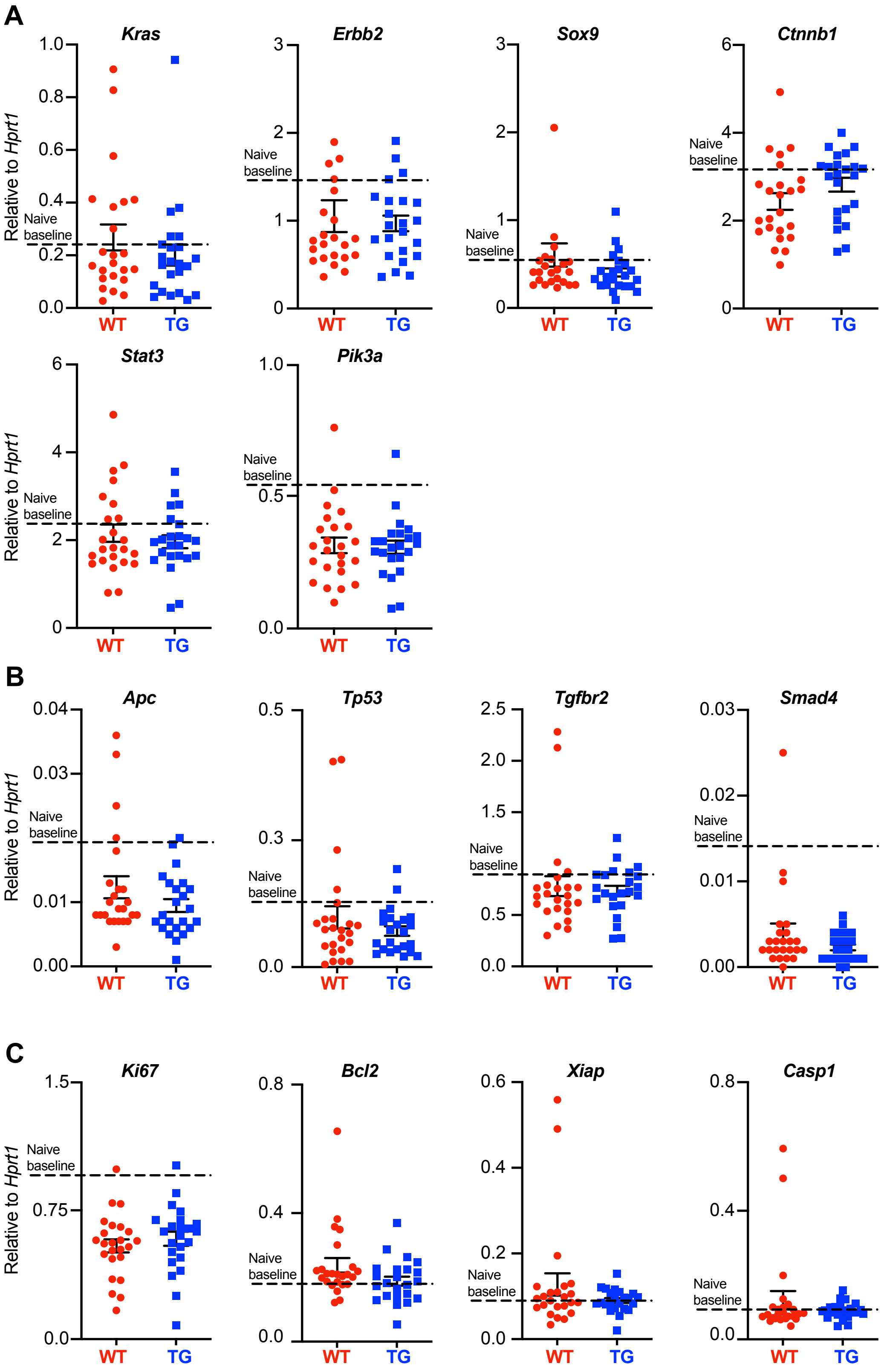
